# Therapeutic Distance: An Orbit-Based Framework for ICU Decision Support Initial Validation in 11,627 Sepsis Patients from MIMIC-IV

**DOI:** 10.64898/2026.04.02.26350049

**Authors:** Alexis Basilakis

## Abstract

**Background:** Patient matching in intensive care databases yields sample sizes too small for individualised outcome analysis. Current AI systems provide population-level guideline summaries but omit stratification variables that may invert therapy signals at the individual level.

**Methods:** We developed the Therapeutic Distance framework, which computes the z-standardised distance between a patient’s clinical parameters and the centroid of MIMIC-IV patients who received each therapy: d(P,T) = Σ w_i_(T) · |(L_i_ − μ_i_(T)) / σ_i_|. We hypothesise that patients at the same distance to a therapy (same orbit) have comparable outcomes. Six validation experiments were performed on 11,627 sepsis patients (SAPS-II 30–80) from MIMIC-IV v3.1.

**Results:** Echo-stratified vasopressin recipients showed mortality of 30.1% (n=146, 95% CI 22.6–37.7%) versus 53.9% without echo (n=2,426, 95% CI 51.9–55.9%). Confidence intervals did not overlap (bootstrap, 1,000 resamples). However, echo-stratified patients had lower general severity (SAPS-II 49.2 vs 53.9) but higher cardiac biomarkers (troponin 1.0 vs 0.51 ng/mL), indicating that the observed difference is compatible with both severity confounding and a possible cardiac-specific vasopressin effect. Leave-one-out prediction with uniform weights achieved AUC 0.61 as a structural baseline.

**Conclusions:** Therapeutic Distance replaces patient matching with orbit matching, substantially increasing usable sample sizes. The echo-vasopressin finding is hypothesis-generating and mechanistically plausible but not causally proven. The framework is intended as a clinical decision support signal under uncertainty, not as a causal inference method.

## Introduction

Individualised treatment decisions in the intensive care unit require integration of patient-specific physiology with published evidence and database-derived outcomes. Three approaches currently exist, each with fundamental limitations.

First, clinical decision support systems based on large language models deliver guideline-level recommendations but systematically omit stratification variables that may determine whether a therapy helps or harms a specific patient.^1,2^ This omission problem has been described as the ‘Goddard Gap’ in prior work.^3^

Second, reinforcement learning approaches, exemplified by the AI Clinician,^4^ learn optimal treatment policies from observational data but produce opaque recommendations that clinicians cannot verify at the bedside.

Third, patient matching (digital twin) approaches attempt to find identical patients in historical databases. For complex ICU patients with multiple comorbidities, this yields sample sizes of n=1 to 5, insufficient for statistical inference.

We propose a different approach: Therapeutic Distance. Rather than matching patients to patients, we compute the distance between a patient and each available therapy in z-standardised clinical parameter space. We hypothesise that two patients who differ in diagnosis, comorbidities, and demographics but have the same distance to a given therapy occupy the same therapeutic orbit and have comparable outcomes. This transforms sample sizes from single-digit patient matches to cohorts of hundreds.

We validate this hypothesis in septic shock using MIMIC-IV, with a primary focus on the vasopressin-echocardiography collision point, where unstratified analysis may cancel opposing subgroup effects.

## Methods

### Data Source and Study Population

We used MIMIC-IV v3.1 (Medical Information Mart for Intensive Care), a publicly available critical care database from Beth Israel Deaconess Medical Center, Boston, containing 546,028 ICU admissions.^5^ The study population comprised adult patients with sepsis or septic shock (ICD-10: A41.x, R65.2x; ICD-9: 995.91, 995.92, 785.52), SAPS-II 30–80, with lactate and creatinine available within 24 hours. After filtering, 11,627 patients were included.

### Clinical Parameters

Seven bedside parameters were selected: lactate (L_1_), creatinine (L_2_), pH (L_3_), PaO_2_/FiO_2_ ratio (L_4_), NT-proBNP (L_5_), troponin (L_6_), and norepinephrine infusion rate (L_7_, mcg/kg/min). For each patient, the mean value within the first 24 hours of ICU admission was extracted. Norepinephrine dose was derived from the rate field in MIMIC-IV inputevents (itemid 221906, unit: mcg/kg/min).

### Therapeutic Distance Formula

For each therapy T, a centroid μ(T) was computed as the mean of each parameter across all MIMIC-IV sepsis patients who received that therapy. The therapeutic distance between patient P and therapy T was defined as:

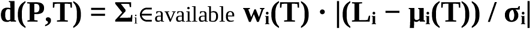

where L_i_ is the patient’s value for parameter i, μ_i_(T) is the therapy centroid, and σ_i_ is the population standard deviation of parameter i across the sepsis cohort. Division by σ_i_ z-standardises each parameter so that variables with different scales contribute proportionally. Therapy-specific weights w_i_(T) allow parameters to contribute differently to each therapy’s orbit; in this initial validation, uniform weights w_i_(T) = 1 were used for all therapies, with z-standardisation providing scale normalisation.

The sum runs only over parameters available for the individual patient. Missing values reduce the dimensionality of the distance computation rather than requiring imputation.

### Orbit Definition and ε-Calibration

Two patients P_1_ and P_2_ are defined as occupying the same therapeutic orbit with respect to therapy T if |d(P_1_, T) − d(P_2_, T)| < ε(T), where ε(T) is a therapy-specific tolerance determined via the enrichment method: the largest orbit radius in which therapy recipients are at least 2-fold overrepresented relative to their baseline prevalence.

#### Important limitation

The orbit hypothesis — that patients at the same therapeutic distance have comparable outcomes regardless of diagnosis — is a modelling assumption. Experiment 1 provides a partial test; formal comparison against propensity-score matching and k-nearest-neighbour approaches is planned.

### Therapy Definitions

Nine therapy nodes were defined from MIMIC-IV prescription, input event, and procedure records within the first 24 hours: vasopressin, echo-guided vasopressin (vasopressin plus echocardiography procedure code), early CRRT, esmolol, furosemide, high-dose norepinephrine (>0.5 mcg/kg/min), prone positioning, sodium bicarbonate, and angiotensin II.

### Validation Experiments

**Experiment 1 — Leave-One-Out Prediction**. 500 patients removed individually; orbit mortality compared to actual outcome (AUC).

**Experiment 2 — Gradient Analysis**. Empirical partial derivatives *∂*M(T)/*∂*L_i_ computed per parameter-therapy pair.

**Experiment 3 — Collision Resolution and Echo Baseline**. Vasopressin mortality compared with and without echocardiography; baseline characteristics compared between groups.

**Experiment 4 — Bootstrap Stability**. Each subgroup resampled 1,000 times; separate 95% CIs computed for echo and non-echo vasopressin recipients.

**Experiment 5 — ε-Calibration**. Orbit width swept across 100 values; enrichment and mortality recorded.

**Experiment 6 — Dimensionality**. Ablation study: each parameter removed one at a time; AUC recomputed. Parameter subsets tested.

### Statistical Analysis and Scope of Inference

All analyses were performed using Python 3.14 with Google BigQuery. Bootstrap CIs used 1,000 resamples (percentile method). Concordance was used for discrimination.

#### We do not interpret mortality differences between therapy groups as causal treatment effects

All mortality comparisons are observational and subject to confounding by indication. The framework is intended as a clinical decision support signal under uncertainty, not as a causal inference method. Propensity-adjusted analysis is required to approximate causal inference and is planned as a next step.

### Ethics

MIMIC-IV is a de-identified, publicly available database accessed through PhysioNet with completed CITI training. No IRB approval was required.

## Results

### Study Population

Of 546,028 ICU admissions, 11,627 met inclusion criteria (sepsis/septic shock, SAPS-II 30–80, lactate and creatinine within 24h). Overall mortality was 34.1%.

### Experiment 4: Bootstrap Stability

### Experiment 3: Collision Resolution and Echo Baseline Comparison

To assess whether the mortality difference reflects a true cardiac-specific effect or severity confounding, we compared baseline characteristics between echo and non-echo vasopressin patients (Table 2).

**Table 1.**
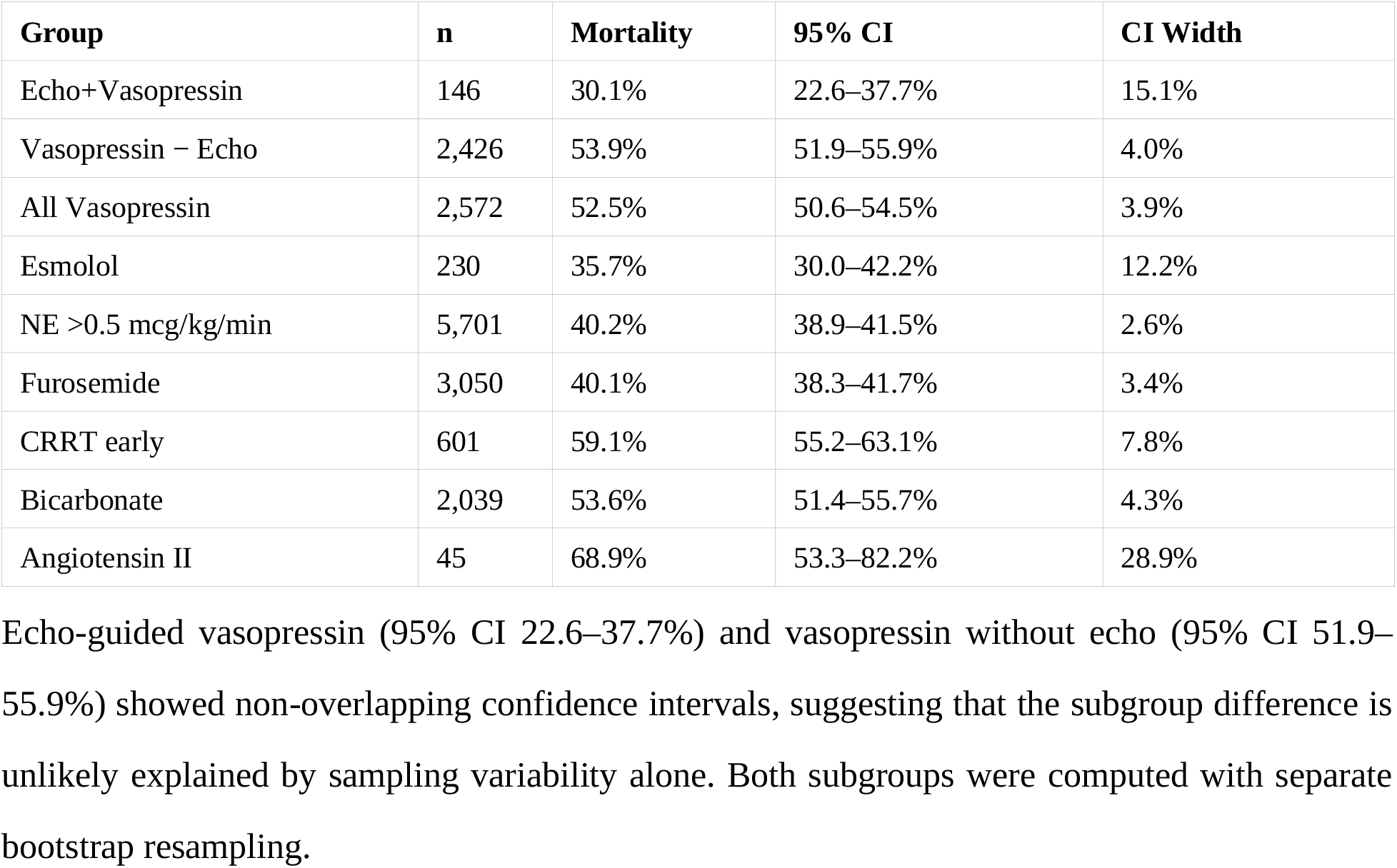
Observed mortality with bootstrap 95% confidence intervals (1,000 resamples) by therapy group and echo stratification. These are observational rates, not causal treatment effects.

**Table 2.** Baseline characteristics of vasopressin recipients stratified by echocardiography. Values are mean ± SD.

| Parameter | Echo (n=230) | No Echo (n=3,018) | Delta |
| --- | --- | --- | --- |
| SAPS-II | 49.2 ± 11.6 | 53.9 ± 12.8 | -4.7 |
| SAPS-II predicted mort. | 44.3% | 53.3% | -9.0% |
| Lactate (mmol/L) | 3.48 ± 2.83 | 4.45 ± 3.62 | -0.97 |
| Creatinine (mg/dL) | 2.21 ± 1.67 | 2.24 ± 1.63 | -0.03 |
| pH | 7.318 ± 0.074 | 7.288 ± 0.096 | +0.030 |
| NT-proBNP (pg/mL) | 11,241 | 10,869 | +372 |
| Troponin (ng/mL) | 1.00 | 0.51 | +0.49 |
| NE rate max (mcg/kg/min) | 0.42 ± 0.64 | 0.50 ± 0.71 | -0.08 |
| NE rate avg (mcg/kg/min) | 0.20 | 0.25 | -0.05 |
| Observed mortality | 27.4% | 53.2% | -25.8% |

Echo-stratified patients had lower general severity (SAPS-II 49.2 vs 53.9, lower lactate, higher pH) but higher cardiac biomarkers (troponin 1.00 vs 0.51 ng/mL). Norepinephrine dosing was comparable between groups (0.42 vs 0.50 mcg/kg/min, max rate). This pattern is consistent with echo being performed in patients with suspected cardiac involvement who are generally less severely shocked but more cardiologically compromised.

The lower SAPS-II predicted mortality in the echo group suggests that part of the observed mortality difference reflects lower baseline severity. The observed difference is fully compatible with baseline severity differences; any residual cardiac-specific effect cannot be separated from confounding in the current observational design. Unmeasured confounders — including differences in care quality, team experience, or institutional practice patterns associated with echo availability — may account for part or all of the residual difference.

This finding is consistent with the neutral results of VASST^6^ and VANISH.^7^ If treatment effect heterogeneity exists along a cardiac axis — with echo-identified cardiomyopathy patients responding differently than non-cardiac patients — then trials that do not stratify by echocardiography would average opposing subgroup effects toward null. This provides a plausible, hypothesis-generating explanation consistent with the neutral outcomes observed. A prospective trial stratifying vasopressin initiation by point-of-care echocardiography would be required to test this hypothesis.

### Experiment 2: Gradient Analysis

Empirical partial derivatives revealed that pH dominated mortality gradients across all therapies (*∂*M/*∂*pH 0.68–2.49), reflecting disease severity rather than therapy-specific signal. Secondary gradients showed therapy-appropriate patterns: troponin ranked second for echo-guided vasopressin, creatinine ranked second for esmolol. The dominance of pH as a severity marker confirms that therapy-specific differential weights are needed for clinical implementation. Current results with uniform weights represent a lower bound on framework performance.

### Experiment 5: ε-Calibration

With uniform weights, therapy enrichment ranged from 0.6× to 2.2× baseline prevalence. Only angiotensin II achieved the ≥2.0× threshold (2.2× at ε=3.30). This confirms that uniform weights are insufficient for therapy-specific orbit definition and that weight optimisation is a prerequisite for clinical implementation.

### Experiment 1: Leave-One-Out Prediction

Leave-one-out prediction on 500 patients (781 patient-therapy predictions) with uniform weights achieved AUC 0.61. Calibration showed correct directionality (higher predicted mortality associated with higher actual mortality) with systematic underestimation consistent with confounding by indication. The objective was not predictive performance but structural validation that orbit-based aggregation carries discriminative signal above chance under unoptimised conditions. The result confirms this but does not represent an optimised prediction model.

### Experiment 6: Dimensionality

Ablation revealed that data completeness, not parameter selection, is the primary constraint. Removing NT-proBNP increased available patients 8-fold (20 to 160). The cardiac-axis subset (5 parameters including proBNP) achieved the highest AUC (0.76, n=20). The renal-axis subset (4 parameters) achieved AUC 0.59 (n=342). No single parameter removal reduced AUC below baseline, indicating the system should compute with variable dimensionality.

## Discussion

This study introduces Therapeutic Distance as a mathematical framework for ICU decision support. Rather than matching patients to patients or learning opaque treatment policies, it computes the distance between a patient and each therapy in z-standardised clinical parameter space. A simple orbit-based distance framework can reveal clinically plausible subgroup structure that standard unstratified analysis obscures.

The central finding is the vasopressin-echocardiography signal: echo-stratified patients showed 30.1% mortality versus 53.9% without echo, with non-overlapping bootstrap confidence intervals. This must be interpreted cautiously. Echo patients were less severely ill by general measures (SAPS-II 49.2 vs 53.9) but more cardiologically compromised (troponin 1.00 vs 0.51). Norepinephrine dosing was comparable (0.42 vs 0.50 mcg/kg/min), suggesting that treatment aggressiveness was similar. The severity difference partially explains the mortality gap; whether the residual difference reflects cardiac-specific vasopressin benefit (V1 receptor selectivity sparing the myocardium from β1-mediated catecholamine toxicity) or unmeasured confounding remains to be determined.

The finding provides a plausible explanation for the neutral results of VASST^6^ and VANISH.^7^ Neither trial stratified by echocardiography. If treatment effect heterogeneity exists along a cardiac axis, population-level analysis would average opposing effects toward null. This hypothesis requires prospective testing.

The concept of therapeutic distance — measuring how far a patient is from a therapy rather than from another patient — is analogous to Kepler’s observation that planetary behaviour depends on orbital distance rather than composition.^8^ The mathematical architecture is structurally generalisable to any clinical space where therapies compete and outcomes are recorded, though it requires domain-specific calibration of parameters, centroids, weights, and orbit tolerances. This study validates the approach in septic shock only.

Current limitations are substantial. Uniform weights fail to separate severity signal (pH) from therapy-specific signal, yielding modest orbit enrichment (0.6–2.2×). AUC 0.61 represents a structural baseline under the simplest possible conditions. Propensity-adjusted analysis is required to approach causal inference. The orbit hypothesis itself requires formal testing against alternative stratification methods.

Despite these limitations, the framework demonstrates that orbit-based stratification reveals clinically plausible subgroup signals hidden in unstratified data, that bootstrap analysis supports the stability of key subgroup differences against sampling variability, and that the architecture accommodates variable dimensionality. These properties make it a candidate for further development as a bedside decision support tool, provided observational limitations are explicitly maintained.

### Limitations

First, all analyses are retrospective and observational from a single US centre (BIDMC, Boston). External validation is required. Second, confounding by indication affects all therapy-outcome associations; propensity-adjusted analysis is planned but not yet performed. Third, uniform weights were used; therapy-specific weights are needed. Fourth, echocardiography is identified via procedure codes, likely underestimating true echo usage; point-of-care echo is frequently undocumented. Fifth, prone positioning (n=2) could not be analysed. Sixth, AUC 0.61 represents a structural baseline. Seventh, the echo group had lower general severity (SAPS-II 49.2 vs 53.9); part of the mortality difference may reflect this imbalance rather than a treatment-specific effect. Eighth, n-values for the echo subgroup differ slightly between analyses (146–230) depending on join completeness with laboratory data; the most conservative estimate (n=146) was used for the primary bootstrap analysis.

## Conclusions

Therapeutic Distance provides an interpretable, structurally generalisable framework for ICU decision support based on orbit matching. Initial validation demonstrates above-chance discrimination and identifies a vasopressin-echocardiography subgroup signal that is hypothesis-generating, mechanistically plausible, and consistent with published trial results, but not causally proven. The framework is intended as a decision support signal under uncertainty. Prospective evaluation, propensity-adjusted analysis, and therapy-specific weight optimisation are required before clinical implementation.

## Data Availability

MIMIC-IV v3.1 is available through PhysioNet. Validation scripts are available at https://chicxulub.ai.

## Acknowledgments

The author thanks PD Dr. Martin Dünser (Kepler University Hospital Linz) for clinical guidance.

## References

1. Hager P, Jungmann F, et al. Evaluation and mitigation of the limitations of large language models in clinical decision-making. Nat Med. 2024;30:1174–1181.

2. Omiye JA, Lester JC, et al. Large language models propagate race-based medicine. NPJ Digit Med. 2023;6:195.

3. Basilakis A. The Goddard Gap: Measuring systematic omissions in AI-assisted clinical decision support. chicxulub.ai. 2026.

4. Komorowski M, Celi LA, Badawi O, Gordon AC, Faisal AA. The Artificial Intelligence Clinician learns optimal treatment strategies for sepsis in intensive care. Nat Med. 2018;24:1716–1720.

5. Johnson AEW, Bulgarelli L, Shen L, et al. MIMIC-IV, a freely accessible electronic health record dataset. Sci Data. 2023;10:1.

6. Russell JA, Walley KR, Singer J, et al. Vasopressin versus norepinephrine infusion in patients with septic shock (VASST). N Engl J Med. 2008;358:877–887.

7. Gordon AC, Mason AJ, Thirunavukkarasu N, et al. Effect of early vasopressin vs norepinephrine on kidney failure in patients with septic shock (VANISH). JAMA. 2016;316:509–518.

8. Kepler J. Harmonices Mundi. 1619.

